# White matter hyperintensities as biomarkers in ALS: link to disease severity, progression, survival, and medication response

**DOI:** 10.64898/2026.04.28.26351973

**Authors:** Katherine Chadwick, Yashar Zeighami, Shima Raessi, Isabelle Lajoie, Canadian ALS Neuroimaging Consortium (CALSNIC), Sanjay Kalra, Mahsa Dadar

## Abstract

**Importance:** While prior work in other neurodegenerative disorders link white matter hyperintensities (WMHs) to disease severity and progression, they remain unexplored in ALS.

**Objective:** To investigate the relationship between presence and progression of WMHs, disease severity, survival, and medication efficacy in ALS.

**Design:** This retrospective study uses data from the Canadian ALS Neuroimaging Consortium (CALSNIC), containing prospectively acquired multicentre longitudinal (three time points over one year) MRI and clinical assessments between 2014 and 2022.

**Setting:** Multicentre study across 9 North American sites.

**Participants:** Participants with a diagnosis of possible, probable, laboratory-supported probable or definite ALS and healthy controls were included. Participants with prior brain trauma were excluded; controls with cognitive impairment or stroke were also excluded.

**Main Outcome(s) and Measure(s):** The main outcomes were differences in baseline and progression of WMHs in ALS patients compared to controls. Secondary outcomes included associations between WMH progression and ALS progression, and subgroup differences (short versus long survival, treatment vs non-treatment groups).

**Results:** Following exclusion criteria, 204 ALS (mean [SD] age, 59.7 [10.4] years; 71 females [34.8%]) and 165 control (mean [SD] age, 55.8 [9.64] years; 70 females [42.8%]) participants were included. ALS patients showed 35.7% greater WMH burden at baseline (p<0.005) and experienced 0.9 cubic centimeters (CCs) more WMH progression over one year (p<0.0001) compared to age and sex matched controls. ALS patients experienced 2 and 4 point drops in ALSFRS-R (p<0.0005) and ECAS-ALS (p<0.005) scores respectively for every 1 CC of WMH progression they experienced. The short survival group (N = 51) experienced faster WMH progression (0.690 CC per year, p<0.05) than the long survival group (N = 75). Patients taking edaravone (N = 181) and riluzole (N = 112) experienced slower WMH progression (0.764 and 0.924 CC per year, respectively, p<0.0005) than those who did not take these medications (N = 23 and N = 90, respectively).

**Conclusions and Relevance:** WMH burden and progression were associated with ALS disease severity, progression, and survival. Edaravone and riluzole treatments were associated with slower WMH progression.

**Key Points:** *Question:* Is the burden of white matter hyperintensities (WMH), and their progression, linked to ALS diagnosis, clinical progression, survival, and medication treatment?

*Finding:* This retrospective study revealed significantly greater WMH burden and progression in ALS compared to healthy controls, as well as links between WMH progression and clinical progression and differences across survival and treatment groups.

*Meaning:* WMHs may be utilized as a biomarker for ALS, and should be integrated into prognostic modeling and clinical trial design.

## Introduction

Amyotrophic Lateral Sclerosis (ALS) is a neurodegenerative disease affecting both upper and lower motor neurons, leading to progressive loss of motor function and muscle weakness. ALS is a highly heterogeneous phenotype, with variability in the site of onset as well as patterns and rates of progression. Survival is also heterogeneous, with most patients experiencing respiratory failure resulting in death 3-4 years after disease onset,^1^ and 5-10% of patients surviving beyond 10 years.^2^ Survival and disease prognosis predictions have historically relied on clinical features such as age, sex, and onset site.^2–5^ While imaging has been traditionally used to rule out ALS mimics such as tumors, recent advances in imaging techniques have allowed for development and integration of imaging biomarkers into diagnostic and prognostic models which can decrease diagnostic delays and aid clinical trial design.^5–9^.^10^ Recent work suggests that imaging markers of white matter (WM) degeneration are associated with disease outcomes, prompting a shift in perspective to include WM pathology in ALS research.^11,12^ Prior studies have investigated WM damage through tract integrity and connectivity (using diffusion tensor imaging), demonstrating significant correlations with disease severity and prognostic utility in ALS.^13–15^

White matter hyperintensities (WMHs) are areas of increased signal observed in T2-weighted and fluid-attenuated magnetic resonance imaging (MRI) and are markers of cerebral small vessel disease and WM damage. WMHs have previously been employed to investigate the role of WM pathology in the context of other neurodegenerative diseases.^16–21^ Their underlying pathology includes demyelination, microglial and endothelial activation, edema, and axonal loss, but varies in terms of location and severity.^22,23^ Compared to age-matched cognitively normal controls, WMHs are more prevalent in individuals with neurodegenerative disorders.^20,24–27^ In Parkinson’s disease, Alzheimer’s disease, and frontotemporal dementia, WMHs increase future gray matter atrophy and cognitive decline^16^. In individuals with mild cognitive impairment, higher baseline WMHs predict conversion to Alzheimer’s dementia.^19,21^ While the presence and role of WMHs have been well established in these disorders, to our knowledge, no large-scale study has yet investigated their potential impact on disease progression and clinical outcomes in ALS.

Edaravone and riluzole are commonly used treatments in ALS. Edaravone is a free radical scavenger that was approved in 2001 in Japan to treat cerebral infarctions, protecting neurons from ischemic damage.^28^ Edaravone has been shown to slow disease progression in ALS and was approved in Canada in 2018 as a treatment for ALS.^29,30^ Riluzole is a neuroprotective drug that inhibits glutamatergic neurotransmission and has been evidenced to increase survival time in ALS.^31,32^ Establishing a link between WMHs and ALS treatments could provide valuable insights into their underlying mechanisms in ALS and highlight the potential for incorporation of WMHs into clinical trial design and disease monitoring.

In this study, leveraging a large longitudinal cohort of ALS patients and matched controls, we examined the prevalence of WMHs in ALS and their associations with disease progression, survival, ALS treatments, and clinical outcomes.

## Methods

### Data

This study utilized longitudinal MRI and clinical data from the Canadian ALS Neuroimaging Consortium (CALSNIC).^33^ Inclusion criteria were a diagnosis of possible, probable, laboratory-supported probable or definite ALS according to the Revised El Escorial Criteria.^34^ T1-weighted and FLAIR MRIs were used to quantify WMHs. MRI protocols were harmonized across centers and vendors. Disease severity was assessed using the Revised ALS Functional Rating Scale (ALSFRS-R), a widely used questionnaire that measures disease severity and disability based on functionality during various daily activities.^35^ Cognitive impairment was assessed using the quantile regression-derived ECAS cutoffs for the Edinburgh Cognitive and Behavioural ALS Screen (ECAS) that account for age and education level.^36,37^ Supplemental materials provides details on ECAS and ALSFRS-R. Participants were excluded if they had brain trauma and history of stroke. Controls with cognitive impairment were also excluded.

### Procedures

MRIs were processed using a validated open access image processing pipeline.^38^ Relevant steps included preprocessing,^39^ stereotaxic registration, co-registration of T1 and FLAIR images, registration of longitudinal visits, and automated WMH segmentation using BISON.^40,41^ All steps were visually assessed (by K.C.) and the cases that had significant artifacts or registration errors were excluded (N = 6). Images with minor segmentation errors were manually corrected (N = 7). Total WMH volumes were extracted in mm^3^ in the standard space to account for head size differences.^42^ To facilitate clinical interpretation, we also report WMHs in cubic centimetres (CCs). Change in WMH volume was calculated by subtracting the baseline WMH volumes from the follow-up measurements. Baseline WMH volumes were log-transformed to normalize their skewed distribution. WMH volume changes were not log-transformed as i) they had a normal distribution and ii) to allow for inclusion of participants that experienced WMH regression (i.e. negative WMH volume change).

### Statistical Analysis

Linear regression models were used to compare baseline WMH volumes between patients and controls, with baseline age and sex as covariates:

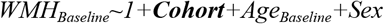

where ***Cohort*** is the categorical variable of interest contrasting patients versus controls. Linear mixed effects models were used to investigate the differences in the rate of WMH progression between patients and controls:

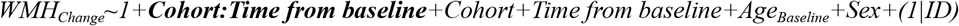

where *WMH Change* indicates the longitudinal change in WMH volume since the baseline visit. *Time from baseline* indicates the time between follow-up and baseline MRIs. In this model, the variable of interest was the interaction term, *Cohort:Time from baseline*, reflecting the differences in the rate of change in WMHs between patients and controls. Participant IDs were included as random effects.

Linear regression and mixed effects models were also used to investigate the associations between baseline WMH burden and WMH progression with baseline disease severity and progression in the patients using ALSFRS-R. Note that the control participants did not complete ALSFRS-R, so these analyses were only performed in the patient group.

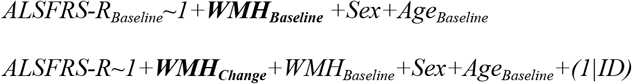

the variable of interest for the cross sectional linear regression model was *WMH*_*Baseline*_ reflecting the overall association between ALSFRS-R and WMH burden, and the variable of interest for the longitudinal model was *WMH*_*Change*_, reflecting the associations between ALSFRS-R scores and change in WMHs, accounting for their baseline levels. Similar models were used to investigate the associations with cognitive changes in patients and controls using ECAS ALS subscore.

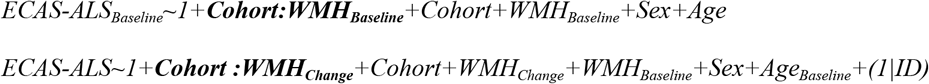

Within patients, similar models were completed to examine differences in three pairs of clinical subgroups: patients with survival data were categorized as “short” or “long” survivors based on their time-to-outcome from the baseline MRI with a 24-month cut-off threshold, and patients were categorized as “edaravone” and/or “riluzole” if they were on these medications at any point in the study period. Similar models were used to contrast cohorts in the patient clinical subgroups, with subgroup instead of cohort as the variable of interest.

## Results

After exclusion of participants based on imaging quality control and clinical criteria, 204 ALS and 165 control participants were included (652 visits). Figure 1 summarizes the participant inclusion process. Table 1 summarizes the characteristics of the participants.

**Table 1.** Characteristics of the participants included in this study. Continuous variables are expressed as mean (SD).

| Group | ALS Patients | Controls | P Value | Short Survival | Long Survival | P Value | Edaravone | Non-edaravone | P Value | Riluzole | Non-riluzole | P Values |
| --- | --- | --- | --- | --- | --- | --- | --- | --- | --- | --- | --- | --- |
| Number | 204 | 165 | N/A | 51 | 75 | N/A | 23 | 181 | N/A | 112 | 90 | N/A |
| Female N (%) | **71 (34.8)** | **70 (42.4)** | p = 0.16 | 16 (31.4) | 29 (38.7) | 0.51 | 9 (39.2) | 62 (34.3) | 0.81 | 40 (35.7) | 31 (34.4) | 0.64 |
| Age | **59.7 (10.4)** | **55.75 (9.64)** | **p <0.001** | 60.2 (8.62) | 57.6 (10.9) | 0.74 | 56.83 (7.22) | 60.09 (10.7) | 0.15 | 59.9 (9.78) | 59.47 (11.1) | 0.76 |
| ALSFRS-R | 37.6 (6.34) | N/A | N/A | **36.47 (6.38)** | **39.02 (5.13)** | **< 0.05** | 38.62 (4.89) | 37.44 (6.49) | 0.42 | 37.3 (6.14) | 37.9 (6.60) | 0.49 |
| ECAS | **105.3 (14.33)** | **114.1 (8.00)** | **p <0.001** | 103.1 (12.2) | 103.4 (14.6) | 0.90 | 109.1 (14.5) | 104.8 (14.3) | 0.18 | 105.3 (13.8) | 105.3 (15.1) | 0.98 |
| ECAS-ALS | **78.04 (11.91)** | **84.79 (6.44)** | **p <0.001** | 76.47 (11.1) | 76.51 (11.4) | 0.98 | 81.32 (9.97) | 77.60 (12.1) | 0.17 | 77.7 (11.4) | 78.46 (12.6) | 0.68 |
| Symptom Duration | 31.68 (31.38) | N/A | N/A | **20.54 (13.7)** | **35.9 (41.9)** | **< 0.05** | 28.14 (13.3) | 32.1 (32.9) | 0.58 | 29.5 (20.9) | 34.6 (41.2) | 0.26 |
| Education | **15.2 (3.51)** | **16.35 (3.20)** | **p <0.001** | **15.65 (3.67)** | **14.38 (2.91)** | **< 0.05** | 15.48 (2.29) | 15.11 (3.63) | 0.64 | 14.95 (3.71) | 15.4 (3.22) | 0.36 |
| Bulbar Onset N (%) | 47 (23.0) | N/A | N/A | **15 (29.4)** | **11 (14.7)** | **0.07** | 2 (8.70) | 45 (24.9) | 0.14 | 23 | 24 | 0.39 |

**Figure 1.**
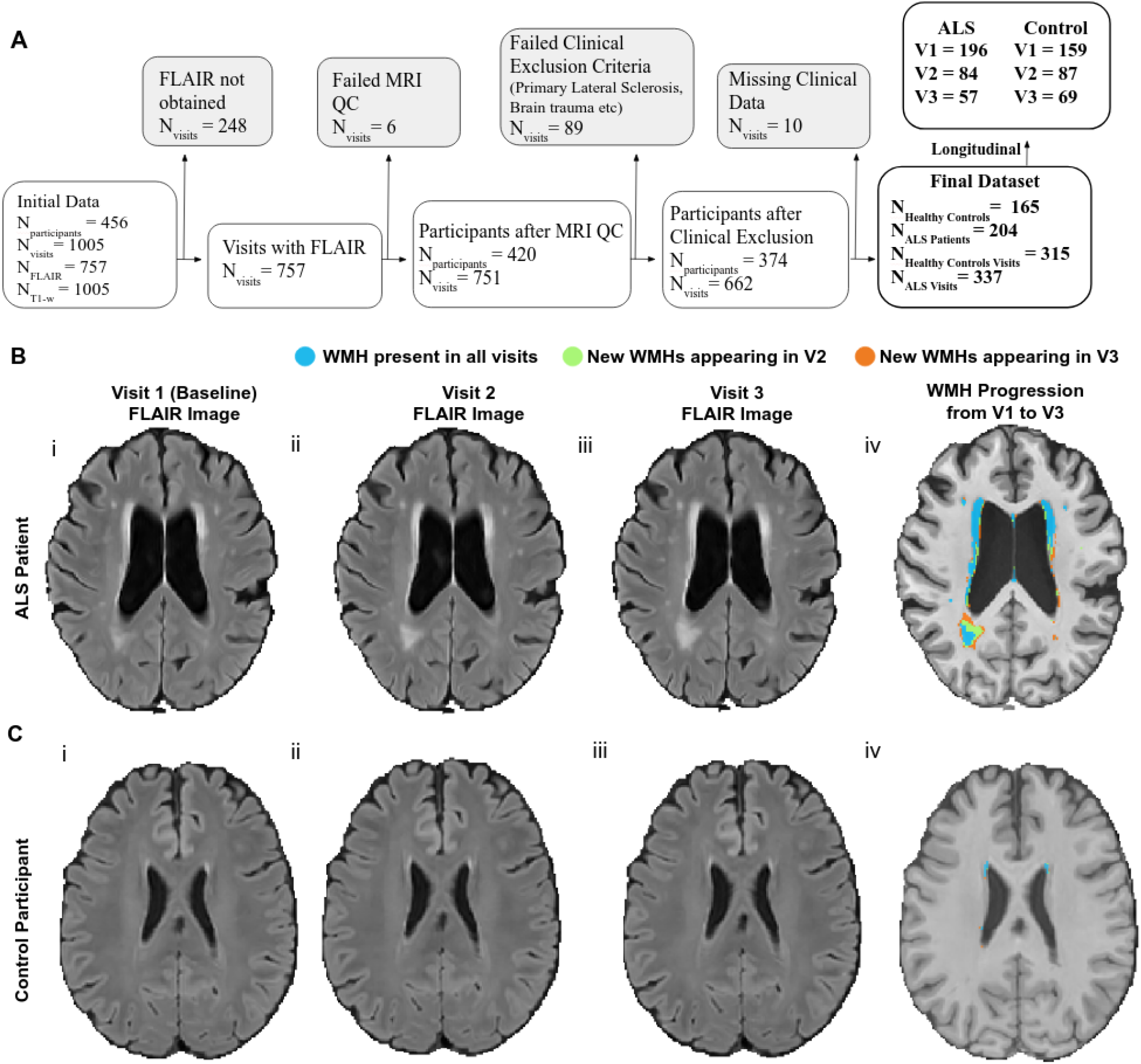
**A) Participant inclusion and exclusion**. MRI exclusions were mainly due to motion artifacts. Clinical exclusions were based on co-presence of other clinical diagnoses and missing variables. **B) i-iii. FLAIR Images of ALS participant** from first timepoint (i) to third timepoint (iii), **iv. WMH progression in ALS Participant;** WMH present in all time points (blue), progress and are present in last two timepoints only (green) and last timepoint only (yellow) **C) i-iii. FLAIR Images of Healthy Control participant** from first timepoint (i) to third timepoint (iii), **iv. WMH progression in Healthy Control participant;** WMH present in all time points (blue), progress and are present in last two timepoints only (green) and last timepoint only (orange)

Accounting for age and sex, baseline WMH volumes were significantly higher in patients than controls (p<0.005) (Table 2, Figure 2.A). Furthermore, WMHs progressed at significantly faster rates in ALS patients compared to controls (over 0.9 CC greater WMH accumulation in one year, p<0.0001) (Table 2, Figure 2.E). ALSFRS-R scores significantly decreased with greater WMH progression in the ALS patients (Table 2, Figure 3.A). For every 1 CC increase in WMH volume, ALSFRS-R scores decreased by 2 (p < 0.001). While baseline WMHs were not differentially associated with cognitive scores across patients and controls, the longitudinal models showed a significant interaction between ALS diagnosis and WMH change, indicating a stronger association between change in WMH and cognition in ALS patients compared to controls (p = 0.001) (Table 2, Figure 3.B).

**Table 2:** Summary results. Values indicate the estimates and p values for the variables of interest. Significance differences and associations are indicated in **bold font**.

|  | Patient vs Control |  | Short vs Long |  | Edaravone vs non-edaravone |  | Riluzole vs non-riluzole |  |
| --- | --- | --- | --- | --- | --- | --- | --- | --- |
| | $\beta$ | P Value | $\beta$ | P Value | $\beta$ | P Value | $\beta$ | P Value |
| Baseline WMH differences<sup>1</sup> | **0.306** | **0.002** | 0.0814 | 0.63 | **-0.546** | **0.001** | -0.0330 | 0.80 |
| WMH change<sup>2</sup> | **901.61** | **< 0.0001** | **689.5** | **0.02** | **-764.17** | **0.0002** | **-924.2** | **< 0.0001** |
| Baseline ALSFRS-R vs WMHs<sup>3</sup> | -0.47 | 0.36 | 0.6361 | 0.50 | -0.9223 | 0.53 | 0.0774 | 0.93 |
| Baseline ECAS-ALS vs WMHs<sup>4</sup> | -1.287 | 0.18 | **-3.0654** | **<0.10** | 3.0586 | 0.27 | -0.9729 | 0.60 |
| ALSFRS-R vs WMH change<sup>5</sup> | **-0.002** | **0.0002** | -0.0008 | 0.47 | **-0.0072** | **0.05** | -0.0006 | 0.60 |
| ECAS-ALS vs WMH change<sup>6</sup> | **-0.004** | **0.001** | -0.0034 | 0.25 | 0.0057 | 0.40 | -0.0006 | 0.77 |

Reported estimates indicate the $\beta$ and P values of the variables of interest in the linear regression and mixed effects models described in Methods section and indicated in **bold font** below:

**Figure 2.**
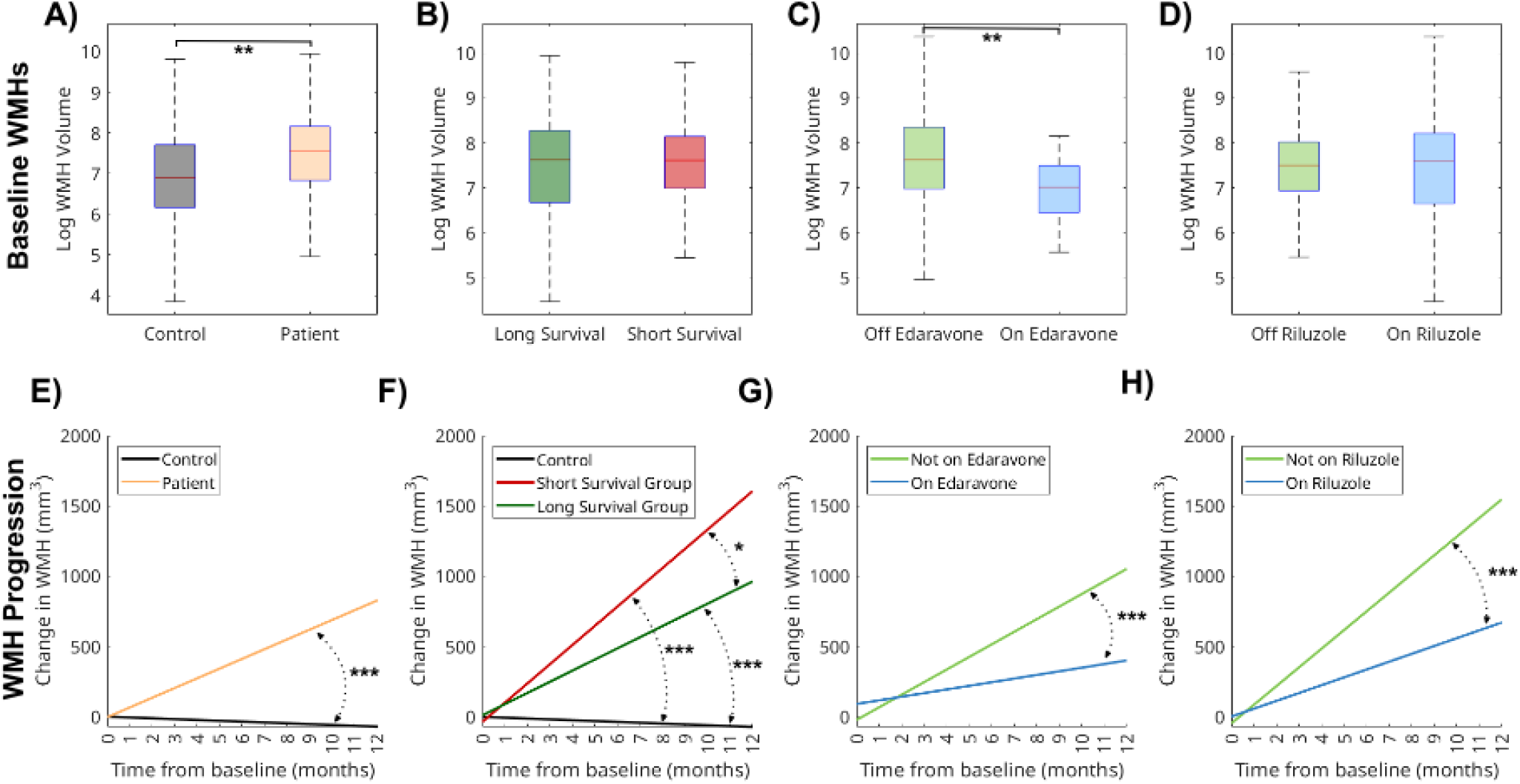
A-D: Log-transformed baseline WMH volumes across **A. patients versus controls:** patients had 35.7% greater WMH burden at baseline (β(log) = 0.3059 p < 0.005), **B. short versus long survival groups:** no significant differences, **C. edaravone versus non-edaravone groups:** patients in the edaravone group had 42.07% greater WMH burden at baseline than those in non-edaravone group (β(log) = -0.546 p < 0.005). **D. riluzole versus non-riluzole groups:** no significant differences, E-F: Predicted progression of WMH in one year across **E. patients versus controls:** patients experienced 0.90 CC more WMH progression (p < 0.0001), **F. short versus long survival groups:** short survivors experienced 0.69 CC more WMH progression (p < 0.05) than the long survival group, **G. edaravone versus non-edaravone groups:** patients in the ‘On’ group experienced 0.76 CC more WMH progression (p = 0.0001), H. **riluzole versus non-riluzole groups:** patients in the ‘Off’ group experienced 0.92 CC more WMH progression (p < 0.0001). Note: Statistical significance reported with asterisks: *p < .05; **p < .01, ***p < .001

**Figure 3.**
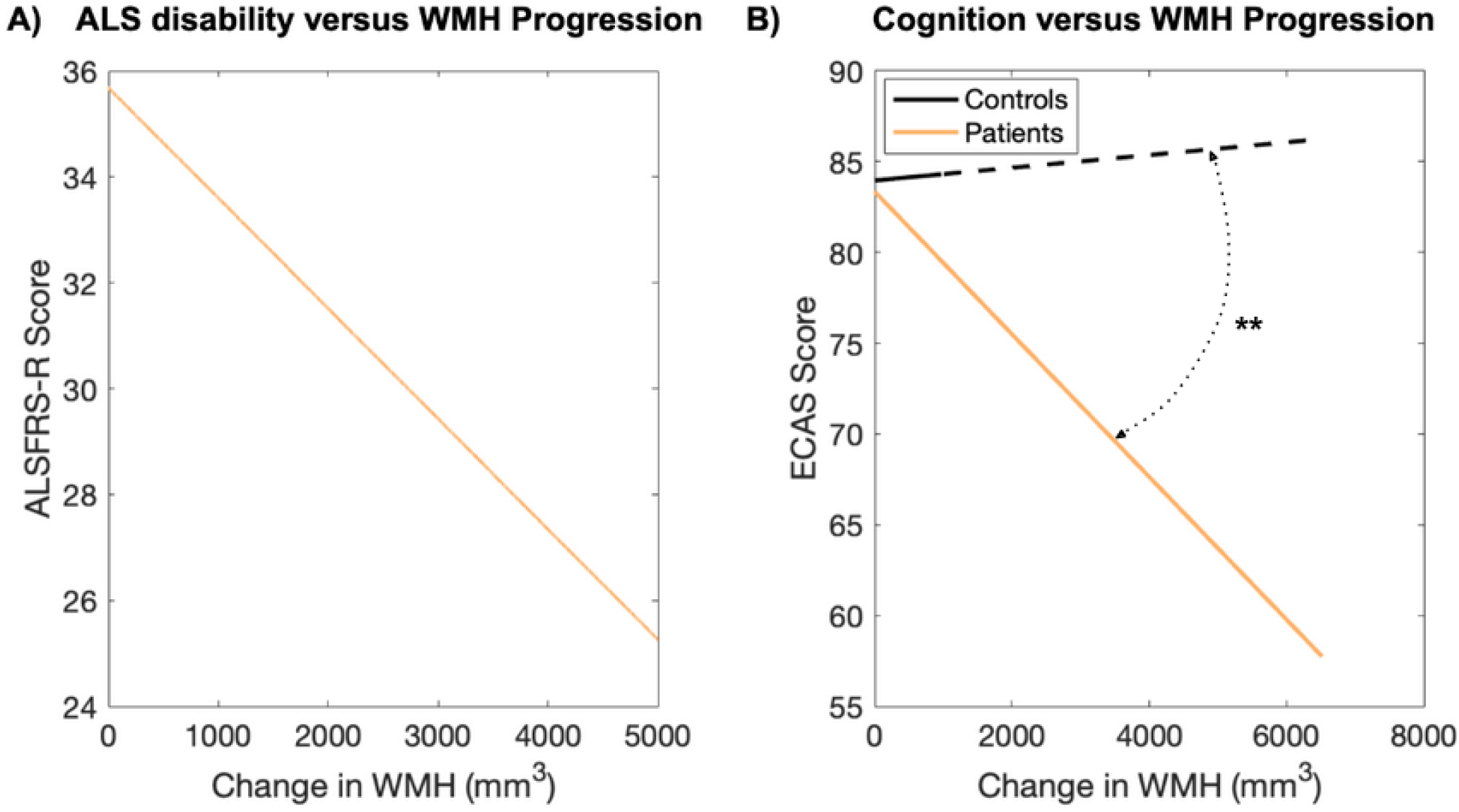
**A. Predicted progression of ALS severity, as measured by ALSFRS-R:** for 1 CC increase in WMH volume, ALFRS-R dropped two full points (p < 0.0005), **B. Predicted progression of cognitive decline, as measured by ECAS-ALS:** for 1 CC increase in WMH volume, ECAS-ALS dropped by 4 full points in the patients compared to controls (p < 0.005). As control participants had a smaller range of WMH progression compared to the patients, extrapolated predictions for the control group beyond their maximum WMH change is depicted as a dashed line.

While there were no baseline differences in WMH volumes between the short and long survival groups, there was a significant interaction between survival group and WMH progression, indicating a faster WMH progression in short survivors (0.690 CC greater WMH accumulation per year, p < 0.05) (Table 2, Figure 2.F). There was a marginally significant interaction between survival group and baseline WMH volume when examining cognition, indicating a marginally stronger association between WMH burden and cognitive deficits in short survivors.

The edaravone group had significantly lower baseline WMH volumes than the non-edaravone group (42.07%; *β*(log) = -0.546 p < 0.005, Table 2, Figure 2.C). There was also a significant interaction between edaravone treatment and WMH progression, indicating faster WMH progression in non-edaravone group (0.764 CC/year, p < 0.001) (Table 2, Figure 2.G). Examining the association between WMH progression and edaravone treatment on clinical measures, there was a small range of WMH progression in the edaravone group (mean [SD] WMH_Change_, 0.175 [0.175] CC). While there were no baseline differences in WMH volumes between the riluzole and non-riluzole groups, there was a significant interaction between riluzole treatment and WMH progression, indicating a faster WMH progression in non-riluzole group (0.924 CC/year, p < 0.0001, Table 2, Figure 2.H).

Reported estimates indicate the β and P values of the variables of interest in the linear regression and mixed effects models described in Methods section and indicated in **bold font** below:

1. Linear regression; *‘WMH*_*Baseline*_ *∼* ***Cohort*** *+ Age*_*Baseline*_ *+ Sex’*. ***Cohort*** contrasts the WMH volumes between groups of interest.
2. Linear mixed effects; *‘WMH*_*Change*_ *∼* ***Cohort:Time from baseline*** *+ Cohort + Time from baseline + Age*_*Baseline*_ *+ Sex + (1*|*ID)’*. ***Cohort:Time from baseline*** indicates the differences in WMH progression slopes between groups of interest.
3. Linear regression; *‘ALSFRS-R*_*Baseline*_ *∼* ***WMH***_***Baseline***_ ***:Cohort*** *+ Baseline WMH + Cohort + Age*_*Baseline*_ *+ Sex’*. ***Baseline WMH:Cohort*** indicates the differences in the associations between baseline WMH volume and ALSFRS-R scores between the groups of interest.
4. Linear regression; *‘ECAS-ALS*_*Baseline*_ *∼* ***WMH***_***Baseline***_***:Cohort*** *+ WMH*_*Baseline*_ *+ Cohort + Age*_*Baseline*_ *+ Sex’*. ***Baseline WMH:Cohort*** indicates the differences in the associations between baseline WMH volume and ECAS-ALS scores between the groups of interest.
5. Linear mixed effects; *‘ALSFRS-R ∼* ***WMH***_***Change***_ ***:Cohort*** *+ WMH*_*Change*_ *+ Cohort + Age*_*Baseline*_ *+ Sex + (1*|*ID)’*. ***WMH Change:Cohort*** indicates the differences in the associations between WMH progression (change in WMH volumes between follow up visits and baseline) and ALSFRS-R scores between the groups of interest. One outlier greater than three standard deviations was excluded from this analysis for the Edaravone cohort.
6. Linear mixed effects; *‘ECAS-ALS ∼* ***WMH***_***Change***_ ***:Cohort*** *+ WMH*_*Change*_ *+ Cohort + Age*_*Baseline*_ *+ Sex + (1*|*ID)’*. ***WMH Change:Cohort*** indicates the differences in the associations between WMH progression (change in WMH volumes between follow up visits and baseline) and ECAS-ALS scores between the groups of interest. One outlier greater than three standard deviations was excluded from this analysis for the Edaravone cohort.

Notes: Baseline WMH volumes were log transformed prior to analyses. WMH Change indicates WMH volume difference between follow up and baseline visits. Baseline visits were not included in these analyses.

CALSNIC included an additional subset of 31 participants that did not have FLAIR images acquired in the protocol. Since BISON can also segment WMHs based on T1w images,^41,43,44^ T1w-based WMHs were quantified for participants that did not have FLAIR data. Supplementary Table S1 summarizes the participants characteristics for the full sample. Tables S1-S3 and Figures S1-S3 summarize the results of the models completed in the full sample (N = 218 patients and N = 182). The results were consistent with the FLAIR based analyses in terms of effect size and significance.

## Discussion

In this study, we investigated the role of WMHs as a biomarker in ALS in a large multi-center cohort of patients and matched controls. Our findings revealed, for the first time, that ALS patients present with significantly greater burden and progression rates of WMHs than matched healthy participants. Furthermore, we demonstrated that WMH burden increases are associated with increased disease progression and disability in ALS. Finally, we showed that treatments targeting neuroinflammation are effective in slowing WMH progression in ALS patients, suggesting a potential underlying mechanism for WMHs in ALS.

The greatest strength of our study is its unprecedented large sample size and multi-site nature, ensuring the generalizability of our findings. The CALSNIC dataset is the largest longitudinal imaging ALS dataset in North America. To our knowledge, this is the first study of this size to investigate WMHs in ALS and observe their relationship with disease progression. Hyperintense signal on T2 or FLAIR, particularly in the precentral gyrus has been observed in ALS.^45–47^ In a sample of 18 ALS patients and matched controls, visually rated subcortical WMHs were present significantly more often in ALS patients.^48^ As well, multiple case reports have described the presence of demyelinating WM lesions in ALS.^49,50^ To our knowledge, our study is the largest of its kind to date. Our WMH segmentation methods have been developed and extensively validated for use in multi-center and multi-scanner studies of aging and neurodegenerative disorders^17,19,41,43^, and provide robust WMH estimations for patient populations.

Our findings demonstrate the potential utility of examining WMHs in ALS. Our longitudinal modeling of WMH progression revealed that patients on average experience over 0.9 CCs per year more WMH progression the controls, adjusting for age, sex, and baseline WMH (Table 2, Figure 2.E1). Furthermore, to be an ideal biomarker, WMHs should also be correlated with disease progression, which our study showed for the first time (Table 2, Figure 3.A). Patient ALSFRS-R scores decreased as WMH increased, with a 1 CC increase in WMH volumes corresponding to a 2 point decrease in ALSFRS-R. In our ALS cohort, WMH volumes increased by 0.28 CCs at the first follow up timepoint, corresponding to 0.56 decrease in ALSFRS-R. At the second follow up, WMH volumes increased by 0.52 CCs, corresponding to a 1.02 point decrease in ALSFRS-R. The ALSFRS-R is a sensitive indicator of quality of life, independence, and survival in ALS. Each point drop in this score leads to a 7% increase in risk for death or tracheostomy.^51^ Slowing the progression of the ALSFRS-R score by 16.5% can indicate increased survival by several months.^52^

There is an urgent need for accurate biomarkers in ALS.^5–9^ ALS diagnosis is dependent on clinical features and leads to an average diagnostic delay of 10 to 16 months after symptom onset.^53^ This limits opportunity for potential neuroprotective treatments.^8,9,53^ WMHs are non-specific to ALS. As such, it is currently difficult to assess the role they may have in ALS diagnosis, monitoring, and prognosis. However, the rate of WMH progression, localisation, and underlying pathology may be unique to ALS and merits future investigation. Uncovering ALS-specific characteristics of WMHs could provide valuable insights to improve diagnostic delay, predict individual disease progression and improve clinical trial design. This study provides novel evidence that WMHs may be used as a sensitive biomarker to indicate disease progression and prognosis. Previous literature has suggested that the integration of imaging biomarkers may improve prognostic models.^10,12,54^ van der Burgh et al. used deep learning to develop a clinical-based, an MRI imaging-based, and an integrated clinical and MRI-imaging based model of disease progression in ALS. Patients classified incorrectly by the unimodal models were often classified correctly when using the integrated clinical and MRI-imaging based model. Future studies developing prognostic models for ALS might benefit from including WMHs as additional features. A recent study utilizing visual rating of corticospinal tract WMH in a smaller subgroup of the 20 patients and 20 controls from CALSNIC found relatively high specificity of corticospinal tract WMHs for ALS, but suggested insufficient sensitivity for diagnostic usage.^55^ However, they did not assess WMH volumes quantitatively or outside of the corticospinal tract. The present work may improve sensitivity for ALS detection through quantitative whole-brain assessment.

There exist limitations in the present work. Due to the inherent challenges in acquiring longitudinal follow-up data in a population with relatively high mortality and disability rates, CALSNIC protocol does not include follow-ups past one year, limiting our ability to examine longer term WMH progression trajectories. In practice, a diagnostic tool for ALS must be able to distinguish between diseases of similar clinical presentation and ALS,^55^ whereas our investigation provided comparison between healthy controls and ALS participants. Characterizing the underlying substrates of WMHs in ALS and their potential differences compared to the WMHs observed in aging and other neurodegenerative disorders warrants further investigations using quantitative MRI sequences with postmortem validations.^44,56,57^ Further, WMH are linked to aging, and our controls were slightly younger than our ALS participants. Our investigation into links between medication treatment and WMH labeled participants as the treatment group if they were on the medication at any point in the study period, which could occlude more subtle WMH changes.

An advantage that WMH offers as a biomarker for ALS is the potential for early interventions. WMHs arise from and are indicators of cerebral small vessel disease, and can be mediated by treatments such as blood pressure controlling medications, preventing the development of further WM hyperintensities. There is emerging evidence that vascular health plays a role in disease progression in ALS.^58,59^ These findings, coupled with our novel evidence of the contribution of WMH to disease progression in ALS, suggest the possibility of vascular risk management, including antihypertensive therapy to slow WMH progression and related ALS progression. Our analyses also revealed slower WMH progression in ALS patients receiving edaravone and/or riluzole treatments. While these are known ALS-specific agents, their association with longitudinal WMH has not been established in the context of ALS. Both drugs target pathways that have links to potential underlying WMH causes. In a rat bilateral carotid artery occlusion model, three-day edaravone treatment increased endothelial protection and BBB barrier, protecting against oxidative damage and WM lesions, and protecting learning memory.^60^ In humans, edaravone has been used to reduce WM damage in ischemic stroke patients.^61^ Riluzole reduces neuroinflammation through decreased excitotoxicity.^32^ In a rat traumatic brain injury model, riluzole treatment reduced vestibulomotor deficits, cerebrovascular permeability, brain edema, and inflammatory markers.^62^ Oxidative stress and neuroinflammation have well-established roles in the progression of ALS,^63^ and our investigation revealed that treatment with edaravone and riluzole may offer a protective role against WMH in ALS, indicating a potential link between oxidative stress and neuroinflammation and as underlying drivers of WMH in ALS. In future studies, neuropathology examinations can offer additional insights into the underlying causes of WMH presence and progression in ALS, and to further investigate potential early intervention targets. Studies are currently investigating the utility of these drugs in other diseases, including Traumatic Brain injuries and Alzheimer’s Disease^61,64^ and in future clinical trials, WMH burden and progression could be a valuable target outcome to evaluate the efficacy of these treatments in the context of ALS.

## Conclusion

This study demonstrated, for the first time, the utility of WMHs as an imaging biomarker for disease diagnosis and prognosis in ALS. ALS patients are significantly more burdened with WMHs than healthy controls. The rate of WMH progression is significantly higher in ALS patients than in controls, and ALS patients with greater WMH burden have significantly worse ALSFRS-R scores, indicating greater disability. As well, the rate of progression of WMH was shown to be significantly increased in ALS patients with short survival times, and participants who were not on riluzole or edaravone medications. Future studies should integrate WMHs into prognostic models to examine their prognostic abilities at the individual patient level.

## Data availability

Imaging and clinical data can be requested from the CALSNIC consortium (https://calsnic.org/).

## Acknowledgements

We are grateful to the CALSNIC study participants, site PIs, research staff and MRI technologists. The authors also acknowledge use of Compute Canada (https://alliancecan.ca/en) resources for performing the image processing analyses in the presented work. See Supplementary Table [S3](https://onlinelibrary.wiley.com/doi/10.1002/ana.27196#support-information-section) for a complete list of the CALSNIC group members. This project was supported by research funds from a ALS-Canada Brain Canada discovery grant. Isabelle Lajoie was also supported by a postdoctoral fellowship from ALS-Canada and Brain Canada Foundation. Katherine Chadwick is also supported by a CIHR CGSM.

## Competing interests

The authors have no competing interests to disclose.

## Notes

### Competing Interest Statement

The authors have declared no competing interest.

### Author Declarations

CALSNIC adheres to the principles of the Declaration of Helsinki and received approval from the Health Research Ethics Boards of all participating sites; list of sites available at https://calsnic.org/our-sites/. The datasets used for this study had been de-identified prior to use in this study.

