## Supplementals for "White matter hyperintensities as biomarkers in ALS: link to disease severity, progression, survival, and medication response"

### Supplementary Materials

#### ALS Disability Score: ALS Functional Rating Scale-Revised

The ALS Functional Rating Scale-Revised (ALSFRS-R) is a validated scale that accurately quantifies ALS related disabilities and represents quality of life.[35] It comprises four main categories of function: bulbar, fine motor, gross motor, and respiratory. Each category has three questions where patients and/or caregivers are asked to place the function on a 0-4 scale from complete dependency to normal function. The ALSFRS-R is scored out of 48 points, with every point loss indicating a decline in quality of life.

#### Cognition: Edinburgh Cognitive Assessment Screen

The Edinburgh Cognitive and Behavioral Assessment Scale (ECAS) was used to examine cognition.[36] This test evaluates 5 main areas of cognition: executive function, fluency, language, memory, and visuospatial. It is scored out of 136 points, with a perfect score being 136 and describing superior cognition. The test is separated into ALS specific and ALS non-specific areas of cognition. The ALS specific criteria is executive function, fluency, and language and is scored out of 100 points. The ALS-nonspecific criteria is memory and visuospatial cognition and is scored out of 36 points. ECAS has been validated to provide an accurate estimation of cognitive impairment, both its domains and severity, in ALS patients, and was designed to minimize the effect of physical disability on cognitive scores.[36]

#### Segmentation Modality

WMHs were segmented based on both FLAIR and T1w images for participants that had both modalities available, and based only on T1w images for a subset of the participants that did not have FLAIR images. The same WMH segmentation pipeline was used for T1w-based WMH segmentations, as it has shown high correlations ($r > 0.9$) between T1w and FLAIR based WMH segmentations.[42] The main analyses presented were completed in the subset of the participants that had FLAIR based WMH segmentations available. The models were further examined in the full sample, including T1w based WMH volumes for the participants that did not have FLAIR images available, as well as an interaction term with a categorical variable reflecting WMH segmentation modality (i.e. contrasting FLAIR versus T1w segmentations) to account for potential differences in FLAIR versus T1w WMH estimations. These following supplemental analyses represent the entire dataset available, utilising T1 and FLAIR imaging-based segmentations where appropriate

**Table S1. Characteristics of the entire dataset of eligible participants, including T1-only participants.** Continuous variables are expressed as mean (SD)

| Group | ALS Patients | Controls | P Value | Short Survival | Long Survival | P Value | Edaravone | Non-edaravone | P Value | Riluzole | Non-Riluzole | P Values |
|---|---|---|---|---|---|---|---|---|---|---|---|---|
| Number | 218 | 182 | n/a | 56 | 81 | n/a | 23 | 186 | | 121 | 97 | |
| Female N (%) | 78(35.7) | 79(43.4) | 0.14 | 19(33..9) | 30(37.0) | 0.85 | 10 (43.5) | 64 (34.4) | 0.53 | 44(36.4) | 34(35.1) | 0.95 |
| Age | 60.16 (10.5) | 55.34 (9.92) | p < 0.00001 | 60.30 (8.90) | 60.15 (11.1) | 0.93 | 57.0 (7.33) | 60.63 (10.7) | 0.12 | 60.06 (9.87) | 60.28 (11.3) | 0.88 |
| ALSFRS-R | 37.67 (6.32) | n/a | n/a | 36.55 (6.36) | 39.26 (5.21) | 0.007 | 38.8 (4.95) | 37.4 (6.52) | 0.36 | 37.5 (6.21) | 37.9 (6.48) | 0.71 |
| ECAS | 105.4 (14.6) | 114.1 (7.97) | p < 0.00001 | 104.0 (11.9) | 103.3 (15.9) | 0.79 | 108.6 (14.6) | 104.6 (14.8) | 0.25 | 105.8 (13.57) | 104.8 (15.9) | 0.65 |
| ECAS-ALS | 78.23 (12.0) | 84.92 (6.35) | p < 0.00001 | 77.19 (10.7) | 76.67 (12.1) | 0.81 | 80.81 (9.92) | 77.66 (12.4) | 0.26 | 78.2 (11.3) | 78.3 (13.0) | 0.99 |
| Symptom Duration | 30.61 (30.6) | n/a | n/a | 19.89 (13.7) | 34.44 (41.0) | 0.012 | 28.2 (13.6) | 31.4 (32.4) | 0.66 | 28.2 (20.3) | 33.68 (39.9) | 0.20 |
| Education | 15.03 (3.52) | 16.32 (3.14) | p < 0.001 | 15.54 (3.58) | 14.30 (3.07) | 0.033 | 15.23 (2.49) | 15.02 (3.68) | 0.79 | 14.93 (3.65) | 15.16 (3.36) | 0.64 |
| Bulbar Onset N (%) | 49(22.5) | n/a | n/a | 16 (28.6) | 11(13.6) | 0.051 | 3 (13.0) | 44 (23.7) | 0.38 | 25(20.7) | 24(24.7) | 0.58 |

**Table S2: Summary of results with T1-based visits included.** Values indicate the estimates and p values for the variables of interest. Significance differences and associations are indicated in **bold font.**

| | Patient vs Control | | Short vs Long | | Edaravone vs non-edaravone | | Riluzole vs Non-riluzole | |
|---|---|---|---|---|---|---|---|---|
| | $\beta$ | P Value | $\beta$ | P Value | $\beta$ | P Value | $\beta$ | P Value |
| Baseline WMH differences[1] | 0.28318 | **0.0030** | 0.1139 | 0.48 | **-0.42706** | **0.003** | 0.1814 | 0.03 |
| WMH change[2] | 403.39 | **6.0513e-05** | **370.2** | **0.099** | **-619.54** | **0.002** | -31.602 | 0.84 |
| Baseline ALSFRS-R vs WMHs[3] | -0.12028 | 0.91 | 0.0045 | 0.996 | -1.3964 | 0.36 | -0.25575 | 0.77 |
| Baseline ECAS-ALS vs WMHs[4] | 0.9128 | 0.33 | -0.3839 | 0.84 | 5.2255 | 0.077 | 0.38882 | 0.81 |
| ALSFRS-R vs WMH change[5] | **-0.002** | **0.0005** | -0.00109 | 0.31 | -0.0074 | 0.10 | -0.0001 | 0.91 |
| ECAS-ALS vs WMH change[6] | **-0.0037** | **0.0033** | -0.00032 | 0.90 | -0.0028 | 0.30 | -0.0007 | 0.74 |

**Table S3: Membership of the Canadian ALS Neuroimaging Consortium (CALSNIC)**

| Name | Title | Affiliation |
|---|---|---|
| Dr. Sanjay Kalra | Principal Investigator | University of Alberta, Edmonton, AB, Canada |
| Dr. Christopher Hanstock | Principal Investigator | University of Alberta, Edmonton, AB, Canada |
| Dr. Alan Wilman | Principal Investigator | University of Alberta, Edmonton, AB, Canada |
| Dr. Dean Eurich | Principal Investigator | University of Alberta, Edmonton, AB, Canada |

| | | |
|---|---|---|
| Dr. Christian Beaulieu | Principal Investigator | University of Alberta, Edmonton, AB, Canada |
| Dr. Yee Hong Yang | Principal Investigator | University of Alberta, Edmonton, AB, Canada |
| Dr. Lawrence Korngut | Principal Investigator | University of Calgary, Calgary, AB, Canada |
| Dr. Richard Frayne | Principal Investigator | University of Calgary, Calgary, AB, Canada |
| Dr. Hannah Briemberg | Principal Investigator | University of British Columbia, Vancouver, BC, Canada |
| Dr. Lorne Zinman | Principal Investigator | University of Toronto, Toronto, ON, Canada |
| Dr. Simon Graham | Principal Investigator | University of Toronto, Toronto, ON, Canada |
| Dr. Angela Genge | Principal Investigator | McGill University, Montreal, QC, Canada |
| Dr. Annie Dionne | Principal Investigator | Université Laval, Quebec City, QC, Canada |
| Dr. Nicolas Dupré | Principal Investigator | Université Laval, Quebec City, QC, Canad |
| Dr. Christen Shoesmith | Principal Investigator | Western University, London, ON, Canada |
| Dr. Michael Benatar | Principal Investigator | University of Miami, Miami, FL, United States |
| Dr. Robert Welsh | Principal Investigator | University of Utah, Salt Lake City, UT, United States |
